# Using an Online Panel to Crosswalk Alternative Measures of Alcohol Use As Fielded in Two National Samples

**DOI:** 10.1101/2023.09.13.23295501

**Authors:** Anna M. Pederson, Scott C Zimmerman, Jacqueline M. Torres, Laura A. Schmidt, Ye Ji Kim, M Maria Glymour

## Abstract

**Introduction:** Accurate estimation of the health effects of drinking is hampered by inconsistent phrasing of questions about alcohol use in commonly-used health surveys (e.g., HRS, NYLS79), and measurement error in brief self-reports of drinking. We fielded an online survey to a diverse pool of respondents, assessing two versions of alcohol use questions. We used the measurement survey responses to evaluate correspondence across question versions and create a crosswalk between versions of alcohol questions from two different nationally representative studies of middle-aged adults. The measurement model can also be used to incorporate measurement error correction.

**Methods:** Respondents to two measurement survey platforms (Centiment and Qualtrics) were asked drinking frequency and quantity questions as phrased in the Health and Retirement Study (HRS: average days per week drank in the last 3 months; quantity consumed on days drank in the last 3 months) and differently phrased questions from the National Longitudinal Survey of Youth 1979 (NLSY79: days drank in last 30 days, average quantity consumed on days drank). The order in which respondents encountered different versions of the questions was randomized. From these questions, we derived measures of average weekly alcohol consumption. In the online panel data, we regressed responses to the HRS question on responses to the NLSY question and vice versa to create imputation models. HRS (n=14,639) and NLSY79 (n=7,069) participants aged 50-59 self-rated their overall health (range 0-4, 0=excellent and 4=poor). NLSY79 or HRS participants’ responses to the alcohol question from the other survey were multiply imputed (k=30) using the measurement model from the measurement survey participant data (k=30). We regressed self-rated health on each alcohol measure and estimated covariate-adjusted coefficients from observed and imputed versions of the questions.

**Results:** The measurement survey (n=2,070) included respondents aged 50+; 64.8% female; 21.4% Hispanic, 23.95% Black, 27.1% White, and 27.6% another (“Other”) self-reported racial/ethnic identity. Associations of observed alcohol question responses with self-reported health were slightly smaller than associations of imputed responses for frequency of alcohol use and consumption on days when alcohol was used. For example, using the HRS version of the frequency of alcohol use (days per week), the estimate for the observed question in HRS respondents was ꞵ =-0.045 [-0.055,-0.036]; and the estimate for the imputed version of the HRS question in NLSY79 respondents was ꞵ=-0.051 [-0.065,-0.037]. The estimated effect of average drinks per week was substantially larger for the imputed version of the measure (ꞵ for the observed question in HRS=-0.002 [-0.004,0.001], ꞵ for the imputed version of the HRS measure in NLSY79 respondents=-0.02 [-0.027,-0.012]). Patterns were similar when using the NLSY79 versions of questions as reported in NLSY79 and imputed for HRS respondents. For example, the estimated effect of average drinks per week was substantially larger for the imputed version of the NLSY79 question (ꞵ for the observed question in NLSY79=-0.006 [-0.01,-0.002], ꞵ for the imputed version of the HRS question in NLSY79 respondents=-0.019 [-0.027,-0.01]).

**Conclusions:** Measurement inconsistencies and imperfect reliability are major challenges in estimating effects of alcohol use on health. Collecting additional data using online panels is a feasible and flexible approach to quantifying measurement differences. This approach may enable measurement error corrections, improve meta-analyses, and promote evidence triangulation.

## Introduction

Alcohol is a high-priority risk factor for healthy aging,^1–3^ but comparing evidence across commonly-used surveys is challenging due to inconsistent measures of alcohol use. Ideally, harmonized gold-standard questions would be asked in all studies. However, in practice, fielded surveys have used a wide variety of questions to assess related dimensions of alcohol use. Questions about alcohol use vary across surveys in many respects, including reference time frame (e.g., drinking in the past 1 week vs 1 month) and whether additional information is provided for reference (e.g., “1 standard drink is 12 ounces of 5% alcohol regular beer, 5 ounces of 12% alcohol wine, or 1.5 ounces of 40% alcohol distilled spirits”). Space on major health surveys is very limited, so multiple versions of questions tapping the same construct are rarely asked in the same survey. As a result, it is difficult to directly combine findings on alcohol use and health from different surveys that used different alcohol assessment questions. This challenge is exacerbated by measurement error in self-reports of alcohol use. Random, additive measurement error will, in expectation, attenuate estimated effects of alcohol use on health, and multiple approaches to correcting the attenuation have been developed^4–7^.

Similar challenges arise for many risk factors in health research, especially behavioral risk factors. One solution is to field a third survey which includes multiple versions of questions and develop a crosswalk between alternative assessments. Because the third data set is being used strictly to develop a measurement model, rather than for more comprehensive research on health, it can be relatively short. Such a measurement survey may be much more efficient and feasible to field than adding new questions to large ongoing, representative surveys.

We evaluated this approach for alcohol frequency questions (Figure 1). We designed and fielded an online panel survey to collect data to allow derivation of a direct crosswalk between measures of alcohol use fielded in the Health and Retirement Study^8^ (HRS) and the National Longitudinal Survey of Youth - 1979 Cohort^9^ (NLSY79). We included similar pairs of questions - one drawn from HRS and one drawn from NLSY79 - on participants’ frequency and quantity of alcohol consumption. We estimated regressions using the online panel survey participants’ responses to the question as phrased in HRS to predict their responses to the alcohol question as phrased in NLSY79; likewise we used the responses to the questions as phrased in NLSY79 to predict responses as phrased in HRS. We used this model to impute how HRS participants would have responded to the alcohol use questions as phrased in NLSY79 and likewise to impute how NLSY79 participants would have responded to the alcohol use questions as phrased in HRS. Finally, in both HRS and NLSY79, we regressed participant self-rated health on alcohol use for each empirical and imputed response to each alcohol question, pooled across imputations.

**Figure 1:**
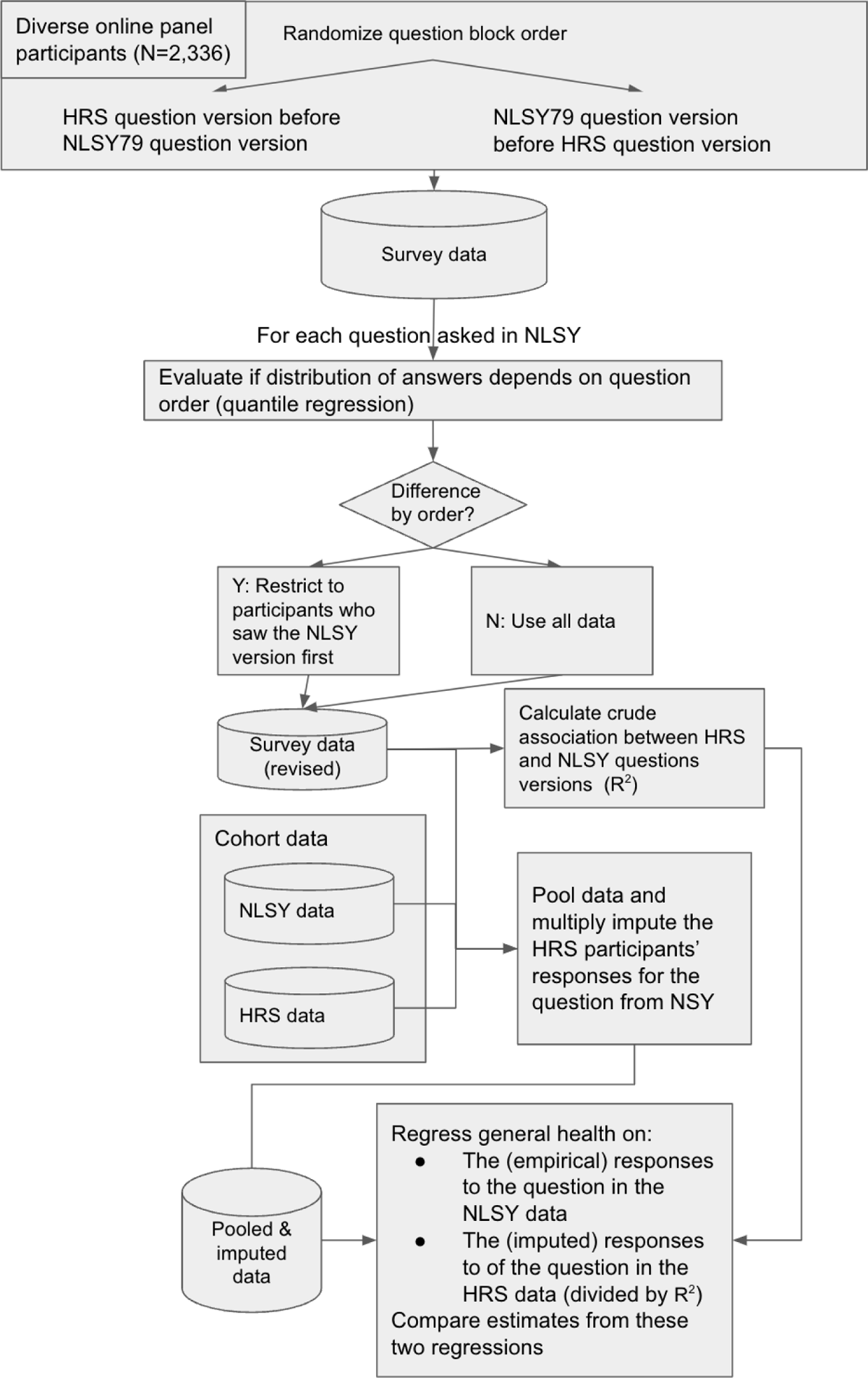
Study design and analysis schematic

## Methods

We designed and fielded a measurement survey using Qualtrics^10^, an online tool used for building and distributing surveys. Respondents were asked 106 questions from seven constructs as phrased in four surveys: the Kaiser Healthy Aging and Diverse Life Experiences (KHANDLE) Study, the Behavioral Risk Factor Surveillance System (BRFSS), the Health and Retirement Study (HRS), and the National Longitudinal Survey of Youth 1979 (NLSY79). The constructs in the survey included questions about alcohol use, smoking status, subjective cognitive status, physical activity, discrimination, activities of daily living, and instrumental activities of daily living. We piloted the survey with a small group of individuals on the research team and others to confirm it was operating as expected.

Two online survey platform companies, Centiment^11^ and Qualtrics, were employed to field the measurement survey to respondents age 50+, aiming to elicit a sample fulfilling the following diversity targets: 25% Asian respondents, 25% Black respondents, 25% Latino/Latina respondents, and 25% White respondents; 33% High school education or less, 33% some college or associate’s degree, and 33% 4-year degree or higher. In this manuscript, we present results from HRS and NLSY79 questions on alcohol use only. Respondents were recruited from two online panels (N=1,272 from Centiment and N=1,063 from Qualtrics) based on the intended sample quotas described above. Centiment and Qualtrics respondents must agree to a Terms of Use and Privacy Policy to participate in surveys. Respondents were compensated upon completion of the survey.

### Centiment data Collection

Centiment recruits respondents to join their research panel through various outlets including Facebook and LinkedIn. Demographic information is collected and reported in panelists’ user profiles. All panelists were pre-recruited and profiled ahead of being targeted for a survey study. Centiment respondents were recruited based on stored profile data and invited to participate via email or SMS. Respondents were compensated upon completion of the survey. Centiment data collection took place from June 23rd 2023 to July 10th 2023.

### Qualtrics data collection

Qualtrics recruits respondents through various sources such as gaming sites, member referrals, and social media. The panels include up-to-date demographic information and respondents were profiled prior to their entry to a survey. Qualtrics respondents were recruited based on demographic characteristics in their user profiles and invited to participate via email invitation, in-app or SMS notifications. Respondents were compensated upon completion of the survey. Qualtrics data collection took place from June 23rd 2023 to August 25th 2023.

### Measurement Survey Design

To ensure good data quality, the survey featured; (1) an attention check (i.e., a survey question that instructed respondents to provide a specific response to the question “We would like to ensure you are reading each question and responding thoughtfully. Please select “Green” as your answer.”); (2) fingerprinting, which looks at IP address, device type, screen size, and cookies to ensure only unique panelists enter the survey; and (3) Captcha verification such as a challenge that the respondent must correctly solve in order to proceed.

The survey asked respondents demographic questions which included age (grouped into seven categories: 50-54; 55-59; 60-64; 65-69; 70-74; 75-84; and 85+); self-reported race/ethnicity (Black, White, Asian, Latino/Latina, and Other), whether or not they spoke a language other than English at home and if so, what language(s); gender (Male, Female, Transgender woman, Transgender man, Two-Spirit, and I use a different term); and educational attainment (Years 0-12 if less than college, Some college but no degree, Associate’s degree, Bachelor’s degree, Master’s degree, and Doctoral or higher level degree). The measurement sample includes people ages 60 and over, although our analytic samples for the national surveys did not. We retained these older individuals to maximize the sample size for the measurement model.

The alcohol questions were copied from the HRS and NLSY79 surveys. The phrasing from HRS was: “In the last three months, on average, how many days per week have you had any alcohol to drink? (For example, beer, wine, or any drink containing liquor)” and “In the last three months, on the days you drink, how many drinks do you have?” The phrasing from NLSY79 was: “During the last 30 days, on how many days did you drink any alcoholic beverages, including beer, wine, or liquor?” and “On the days that you drink, about how many drinks do you have on the average day? By a drink, we mean the equivalent of a can of beer, a glass or wine, or a shot glass of hard liquor”. A response for each alcohol question was required from all respondents.

The survey included a separate block for each survey from which question formats were drawn. We used the randomizer tool in Qualtrics to randomize the order in which each participant saw the blocks.^12^ For 964 respondents, the blocks were time stamped so we were able to establish whether respondents first encountered the HRS version of the alcohol use questions or the NLSY version of the questions. Time-stamp was erroneously not recorded for the remainder of the sample.

### National outcome samples

We applied responses from the online measurement survey to two national surveys: HRS and NLSY79. The HRS is a nationally representative cohort of US adults aged 50+ and their spouses. Every 6 years new enrollments add birth cohorts that have “aged in” to eligibility. Participants are followed up every 2 years. Alcohol frequency and quantity questions were asked starting wave 3 of HRS, so we used participant’s first response from waves 3-15 of HRS, with coding from the RAND public use data file. The NLSY79 is a prospective cohort launched in 1979 with a nationally representative sample of men and women born between 1957 and 1964 who were aged 14 to 22 at baseline.

For comparable age distributions we used data from NLSY79 collected in 2014. The oldest participants from NLSY for which alcohol data were available was 59, thus we limited the samples for both cohorts to adults age 50-59. In addition to the demographic and alcohol questions used to inform the measurement survey, HRS and NLSY79 participants were also asked about self-rated general health (range 0=excellent to 4=poor).

### Data cleaning

To harmonize covariate data between the measurement survey, HRS, and NLSY79, education was coded as less than high school (11 or fewer years of schooling), high school but not a Bachelor’s degree (12 or more years of schooling, or an Associate’s degree or some college reported) or Bachelor’s or more education. The measurement survey included self-reported race/ethnicity categories for “White”, “Black”, “Asian”, “Latino/Latina”, and “Other”, whereas the NLSY79 and HRS data only included race/ethnicity categories “Black”, “White”, “Hispanic”, and “Other”. Therefore, we created a new race/ethnicity variable to reflect the categories in HRS and NLSY79 by grouping “Asian” with “Other”. We classified measurement survey respondents based on their self-identified gender as male or female (regardless of cis- or trans-identity) and grouped the single individual who endorsed being ‘two-spirit’ with male respondents rather than drop that individual from the analyses.

We standardized the HRS and NLSY79 frequency questions to the same scale (days drank per week) by dividing the NLSY79 question by 30 and then multiplying by 7. In addition to days drank per week and drinks per day on days drank, we calculated average weekly drinks as the product of alcohol consumption frequency (days drank per week) and drinks per day on days when alcohol was consumed.

Initial data checks in NLSY and HRS revealed that implausible alcohol quantity values were present in the response data for a small number of respondents. Thus we capped values at the maximum plausible value of 16 drinks/day, affecting 13 HRS and 5 NLSY participants. Our data cleaning protocol included omitting respondents who did not answer the attention check question correctly, but all respondents answered it correctly.

### Analysis

#### Evaluation of order dependency

For the subset of data in which the order of questions presented to the participants was recorded, we created an indicator variable for which question in the pair was answered first. We then assessed whether the distribution of each alcohol question differed depending on whether the participant was randomly assigned to see the NLSY79 version or the HRS version of the question first. We first estimated linear regressions to evaluate whether the mean response differed by question order; we then estimated quantile regressions ^13,14^, predicting each decile of each alcohol measure/quantity with an indicator for whether the respondent encountered the NLSY79 version first. For a given question type (eg. frequency in HRS vs NLSY79), a difference in the distributions by ordering of questions would indicate that separate crosswalks should be derived depending on which question was answered first, while no difference would indicate that a single crosswalk could be derived.

#### Estimation of the measurement model

This model is premised on the assumption that both versions of the alcohol questions are indicators of a latent value of alcohol consumption (i.e., there is a true quantity of alcohol consumption which influences measured alcohol consumption but is subject to random error). In the measurement survey data, we estimated linear regressions for each alcohol variable pair (frequency of drinking; quantity consumed on days drank; and average weekly drinks consumed), regressing the response to the HRS version of the question on the response to the NLSY79 version. For each question pair we calculated crude models adjusting only for the source variable and models with covariates: age group, gender, self-reported race/ethnicity, and educational attainment.

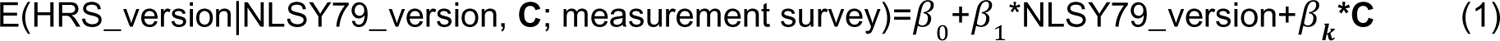

We likewise estimated regressions using NLSY79 versions as the dependent variable and the HRS version as the independent variable.

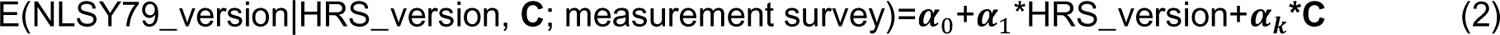

These imputation models were then applied in the outcome data sets, HRS and NLSY79.

#### Application to NLSY and HRS

For NLSY79 participants, we used multiple imputation based on the models estimated in the measurement survey data to estimate their likely responses to each of the alcohol questions as phrased in HRS (HRS_version_imputed). We imputed responses using multivariate imputation by chained equations, generating 30 imputed datasets. We then estimated linear regressions of self-reported health on each alcohol question as asked in HRS and on imputed versions of the questions estimated for each NLSY79 respondent, using equation (1) estimates for the imputation.

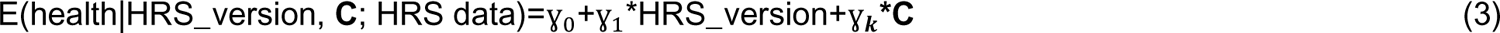

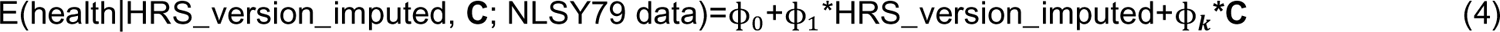

For the imputed versions of the questions, we combined the coefficients ɸ across imputations and estimated standard errors according to Rubin’s rules.^15^ Models were adjusted for sex (Male/Female), race/ethnicity (White, Black, Hispanic, Other), age group (50-54, 55-59), and educational attainment.

The ɣ_1_ and ɸ_1_ estimates are expected to diverge for several reasons. If the alcohol questions are measured with error, we expect ɣ_1_ to be diluted by this error compared to the true effect of alcohol use on health. The ɸ_1_ coefficient on the imputed version of the measure in expectation corrects for this measurement error and thus would typically be larger than ɣ_1_. The corrected estimates may also diverge due to chance in finite samples in HRS and NLSY79 or uncertainty in the estimated crosswalk from the measurement sample. Finally, although HRS and NLSY79 are both national samples of middle-aged adults, in the current analyses we did not apply sampling weights and the samples and the target samples are not identical. For example, NLSY79 participants have remained active in the cohort for over 35 years, whereas HRS participants were recruited much more recently. The actual association of alcohol with health may thus diverge between the populations represented by these two cohorts.

We repeated the imputation and estimation procedure for the NLSY79 versions of the questions.

Analysis was conducted using R version 4.2.2^16^ with the MICE package version 3.16.0^17^ for multiple imputation. IRB exempt approval for this study was granted by the University of California San Francisco IRB with IRB #: 23-38893.

## Results

Descriptive statistics for participants from the measurement survey, HRS and NLSY79 samples are presented in Table 1.

**Table 1:** Sample characteristics.

|  |  | n (proportion) or mean (sd) |  |  |
| --- | --- | --- | --- | --- |
| Variable | Category (if applicable) | HRS | NLSY79 | Online Survey Panel |
| Sociodemographics |  |  |  |  |
| n |  | 14639 | 7069 | 2070 |
| Female |  | 7505 (51.3%) | 3664 (51.8%) | 1341 (64.8%) |
| Race | Black | 3639 (24.9%) | 2165 (30.6%) | 494 (23.9%) |
|  | Hispanic | 2487 (17%) | 1225 (17.3%) | 443 (21.4%) |
|  | Other | 894 (6.1%) | 753 (10.7%) | 572 (27.6%) |
|  | White | 7619 (52%) | 2926 (41.4%) | 561 (27.1%) |
| Education | Less than high school | 2554 (17.4%) | 1112 (15.7%) | 111 (5.4%) |
|  | High school and some higher education | 8050 (55%) | 4681 (66.2%) | 1166 (56.3%) |
|  | Bachelor's degree or more | 3939 (26.9%) | 1276 (18.1%) | 765 (37%) |
| Age group (years) | 50-54 | 9736 (66.5%) | 4611 (65.2%) | 424 (20.5%) |
|  | 55-59 | 4903 (33.5%) | 2458 (34.8%) | 327 (15.8%) |
|  | 60-64 | 0 (0%) | 0 (0%) | 311 (15%) |
|  | 65-69 | 0 (0%) | 0 (0%) | 446 (21.5%) |
|  | 70-74 | 0 (0%) | 0 (0%) | 345 (16.7%) |
|  | 75-84 | 0 (0%) | 0 (0%) | 189 (9.1%) |
|  | 85+ | 0 (0%) | 0 (0%) | 28 (1.4%) |
| Alcohol Use |  |  |  |  |
| Alcohol frequency (days in last 30 days) |  | NA | 1.11 (1.82) | 0.74 (1.35) |
| Alcohol frequency (days per week during last 3 months) |  | 1.23 (1.92) | NA | 1.07 (1.67) |
| Alcohol quantity (average on days consumed) |  | NA | 1.29 (1.71) | 1.06 (1.38) |
| Alcohol quantity (on days consumed in last 3 months) |  | 1.22 (1.97) | NA | 1.12 (1.58) |
| Average weekly consumption (last 30 days) [frequency*quantity] |  | NA | 2.9 (6.8) | 1.41 (2.76) |
| Average weekly consumption (3 months) [frequency*quantity] |  | 3.57 (8.19) | NA | 2.2 (3.71) |
| General Health |  |  |  |  |
| Self-reported general health (Likert scale range: 0=very good to 4=poor) |  | 1.81 (1.16) | 1.62 (1.06) | NA |

### Associations between responses for similar questions

In the measurement survey, responses to HRS and NLSY79 versions of the questions on alcohol frequency and average quantity on days alcohol was consumed were moderately correlated for each variable (frequency R^2^ = 0.60, quantity R^2^ = 0.46), although there were some clear outliers that likely represent misunderstandings of the question. The two versions of the average weekly alcohol consumption variables were more weakly associated (R^2^ = 0.30). See Appendix 3.

### Analytic sample from the measurement survey

Our measurement sample consisted of 2,070 participants. Of the sample, 64.8% of respondents identified as female, approximately 23.9% identified as Black, 21.4% identified as Hispanic, 27.1% identified as White, and 27.6% identified as another racial/ethnic group (“Other”). Respondents self-reported an average of 0.74 (SD = 1.35) days drinking per week in the last 30 days as asked by the NLSY79 version, and an average of 1.07 (SD = 1.67) days drinking per week in the last 3 months as asked by the HRS version. Similarly, respondents reported they averaged 0.74 drinks on days they drank when asked the NLSY79 question version but 1.11 drinks when asked the HRS version. In combination, among measurement sample respondents, the weekly average consumption elicited by the HRS question (2.20 drinks) was notably higher than the weekly average consumption elicited by the NLSY question(1.41 drinks).

### Evaluation of sensitivity of responses to ordering of similar questions

Results from the quantile regressions were used to compare the distribution of alcohol question responses by whether the NLSY block or the HRS block was shown first. These regressions showed no clear difference across quantiles based on the question order (see Figure 2). Therefore, we did not restrict our sample based on the order of the questions.

**Figure 2:**
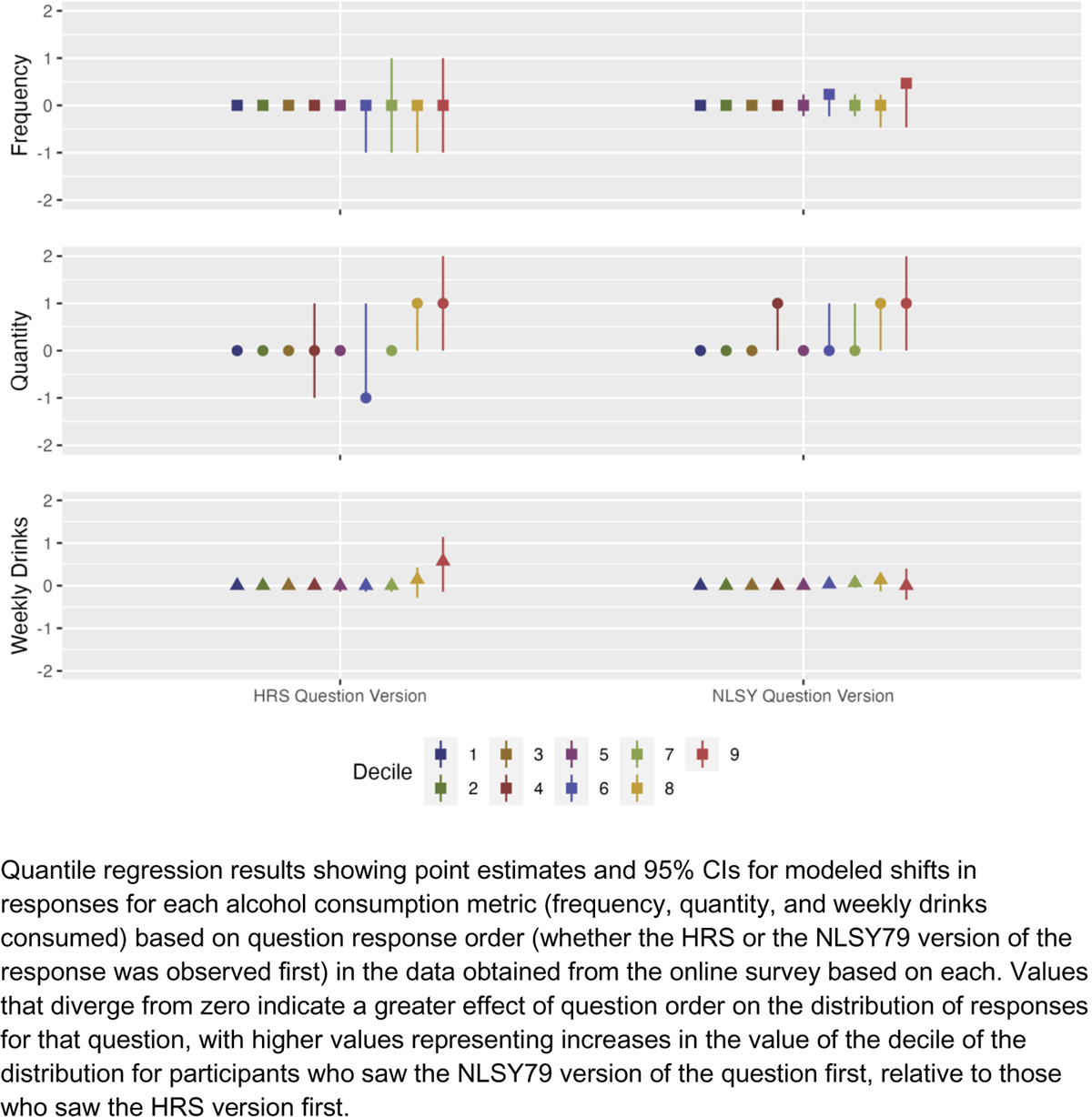
Quantile regression results

### Association of alcohol question responses in the measurement survey

The NLSY79 alcohol frequency question response strongly predicted the HRS alcohol frequency response (b=0.94 [0.90, 0.97]) and likewise the HRS alcohol frequency question response strongly predicted the NLSY79 alcohol frequency response (b=0.65 [0.63, 0.67]). Similar patterns were observed for quantity consumed on days drank (b=0.84 [0.80, 0.87] for NLSY79 version predicting the HRS version and b=0.55 [0.52, 0.57] for HRS predicting NLSY79) and average daily consumption (b=0.95 [0.91, 0.99] for NLSY79 predicting HRS and b=0.53 [0.51, 0.55] for HRS predicting NLSY79 responses).

### Application: Associations of alcohol variables with self-rated health

Results for regressions of self-rated health on each alcohol variable as observed in the data set where it was asked and as imputed in the other data set are shown in Table 2 and Appendix Table 4. Using the HRS versions of the alcohol questions, coefficients for average drinks per week were smaller using the reported version of the question among HRS respondents (b=-0.002 [-0.004,0.001]) than when using the imputed version of the question among NLSY79 participants (b=-0.02 [-0.027,-0.012]). Likewise, coefficients for alcohol use frequency were slightly smaller in HRS (b= −0.045 [-0.055,-0.036]) than for imputed responses in NLSY (b=-0.051 [-0.065,-0.037]). Coefficients for alcohol quantity on days when drinking were also smaller in HRS (b=-0.02 [-0.029,-0.01]) and imputed responses in NLSY (b=-0.043 [-0.06,-0.026]).

**Table 2:** Regression coefficients for general health models.

|  |  | Coefficients<br>Estimate [95%CI], p-value |  |
| --- | --- | --- | --- |
| Domain | Alcohol exposure question version | HRS | NLSY |
| Frequency | "In the last three months, on average, how many days per week have you had any alcohol to drink? (For example, beer, wine, or any drink containing liquor)" (HRS question) | -0.045 [-0.055,-0.036]<br>p<0.01 | -0.051 [-0.065,-0.037]<br>p<0.01<br>(imputed) |
|  | "During the last 30 days, on how many days did you drink any alcoholic beverages, including beer, wine, or liquor? Please enter a numerical value.", times 7/30 (NLSY question) | -0.042 [-0.055,-0.029]<br>p<0.01<br>(imputed) | -0.062 [-0.075,-0.049]<br>p<0.01 |
| Quantity | In the last three months, on the days you drink, about how many drinks do you have? Please enter a numerical value. (HRS question) | -0.02 [-0.029,-0.01]<br>p<0.01 | -0.043 [-0.06,-0.026]<br>p<0.01<br>(imputed) |
|  | On the days that you drink, about how many drinks do you have on the average day? By a drink, we mean the equivalent of a can of beer, a glass of wine, or a shot glass of hard liquor. Please enter a numerical value. (NLSY question) | -0.03 [-0.049,-0.012]<br>p<0.01<br>(imputed) | -0.041 [-0.055,-0.026]<br>p<0.01 |
| Average drinks per week | Product of HRS frequency and alcohol questions: Drinks per week | -0.002 [-0.004,0.001]<br>p=0.15 | -0.02 [-0.027,-0.012]<br>p<0.01<br>(imputed) |
|  | Product of NLSY frequency and alcohol questions: Drinks per week | -0.019 [-0.027,-0.01]<br>p<0.01<br>(imputed) | -0.006 [-0.01,-0.002]<br>p<0.01 |

Each row presents the coefficient, 95%CI and p-value for the coefficient on the alcohol exposure variable for a pair of regressions: one conducted on empirical data and one conducted on imputed data based on the cross-walk. HRS alcohol responses were imputed for questions from NLSY and vice-versa, indicated by "(imputed)".

Using the NLSY versions of the alcohol questions, coefficients for average drinks per week were smaller in NLSY (b=-0.006 [-0.01,-0.002]) than for imputed responses in HRS (b=-0.019 [−0.027,-0.01]). Coefficients for alcohol frequency were larger in NLSY (b=-0.062 [-0.075,-0.049]) than when using the imputed responses in HRS (b=-0.042 [-0.055,-0.029]). Coefficients for alcohol quantity were also slightly larger when using NLSY (b=-0.041 [-0.055,-0.026]) than when using the imputed responses in HRS respondents (b=-0.03 [-0.049,-0.012]).

## Discussion

In this study we collected data from a diverse measurement sample who were asked the HRS and NLSY79 versions of items capturing the frequency and quantity of alcohol consumption. These responses were used to calculate average weekly alcohol consumption for the HRS and NLSY79 question versions. We then examined correspondence between each pair of question versions. We used the observed relationships between questions in the measurement sample to impute values for each alcohol use question as phrased in HRS among respondents to the NLSY79 survey. Likewise, we imputed values for each alcohol use question as phrased in NLSY79 among respondents to the HRS survey. We regressed self-rated health on each observed and imputed alcohol measure in HRS and NLSY79. Estimates for the association between weekly alcohol consumption and self-rated health were similar in the HRS and NLSY79 when using the imputed versions of the alcohol consumption questions and substantially larger than estimates using the non-imputed alcohol use questions in each data set. The analyses demonstrates the feasibility and importance of collecting a measurement sample to crosswalk across commonly used studies of alcohol and health.

The newly collected measurement sample data reflected substantial racial/ethnic diversity by design. The measurement sample respondents reported somewhat lower use of alcohol than HRS orNLSY79 respondents, and were also (by design) slightly older. Differences in response distributions for pairs of similar questions were not sensitive to question ordering for online panel participants: question order effects (e.g. first orienting participants to a 30 day time frame, with subsequent questions re-orienting participants to consider weekly drinking over a 3 month time frame, or vice versa) were not influential on how respondents answered these alcohol questions. Associations between versions of each alcohol measure were moderate in the measurement sample, consistent with substantial potential for measurement error. One possible explanation for inconsistency is that asking people to remember their drinking behavior over different time frames may elicit different responses, even after adjustment for the stated time frame. We observed this phenomenon in our online survey responses (see Appendix 3), for which all HRS versions of the questions (with a 3-month time-frame) resulted in higher estimates of drinking than NLSY versions (which had a 30-day time-frame).

Increased alcohol frequency, quantity, and average weekly consumption were nominally associated with better self-reported health in both NLSY and HRS. This was observed whether using the empirically observed or the imputed version of the metric, possibly capturing the healthy drinker phenomenon.^18–21^ Using the imputed versions of the weekly alcohol consumption variable, estimates were similar in the HRS and NLSY79 samples.

The most important limitation in our approach is that relying on the measurement survey respondents to create a crosswalk requires the assumption that the relationship between two measures is insensitive to features of the survey, including mode of interview and respondent characteristics^23^. It is not feasible to field an interview-based survey instrument that would be faithful to the original studies’ survey modalities: the HRS and NLSY79 survey instruments required long interviews involving a multitude of questions on health and behavior. Completing a crosswalk between these two studies using the original survey modality would involve repeating both surveys for each participant. A further limitation of the current analysis is that we compared only questions as fielded in the HRS and NLSY79 studies; further work is needed to extend these crosswalks to questions fielded in other cohorts, which may yield different results. We used linear models to impute alcohol use questions which are not continuous or even approximately so. Fourth, we studied cross-sectional associations between alcohol consumption and health, and patterns may differ longitudinally. We make no claims here about causal inferences regarding alcohol and health; the goal of this paper is to explore the feasibility of the data collection approach to improving measurement.

This study has several strengths. First, use of an online survey comparing questions as fielded in different surveys is a cost-efficient and very rapid way to gather information to allow for measurement error modeling. With good measurement error models, it is possible to correct for measurement error in other data sets and pool findings across multiple studies that used slightly different versions of the assessments. This allows us to maximize the utility of existing studies by allowing crosswalking between similar questions. This form of data pooling^22^ has the potential to improve estimates from beyond what is possible with a single study alone. Randomizing the question ordering allowed us to assess question order bias. The diverse sample of the online cohort allows us to apply the crosswalk to nationally-representative samples. In future research, we will evaluate specific potential demographic modifiers of the link between the two alcohol measures.

Our study demonstrates the feasibility and value of adding rapidly fielded measurement studies to supplement analyses of major surveillance studies when exposures of interest are measured inconsistently or with imperfect reliability. The viability of online panel studies has increased rapidly. Although these samples may have important limitations for outcome modeling, they are extremely appealing for modeling measurement reliability. Widespread adoption of ancillary measurement studies might be of substantial value in research on health behaviors such as alcohol use. Several research questions remain, including optimal approaches to ensuring validity of online panel responses, preferred statistical approaches for variables with non-normal distributions, and stability of results across data sets and sample composition.

Quantile regression results showing point estimates and 95% CIs for modeled shifts in responses for each alcohol consumption metric (frequency, quantity, and weekly drinks consumed) based on question response order (whether the HRS or the NLSY79 version of the response was observed first) in the data obtained from the online survey based on each. Values that diverge from zero indicate a greater effect of question order on the distribution of responses for that question, with higher values representing increases in the value of the decile of the distribution for participants who saw the NLSY79 version of the question first, relative to those who saw the HRS version first.

## Data Availability

All outcome data produced are available online to registered researchers on the Health and Retirement Study website under data products, and available on the NLS investigator site.

## Appendix

## Appendix 1: Data dictionary

| Category | Variable | Question | Type |
| --- | --- | --- | --- |
| Alcohol frequency | ALC_FREQ_H | In the last three months, on average, how many days per week have you had any alcohol to drink? (For example, beer, wine, or any drink containing liquor) | Continuous |
| Alcohol frequency | ALC_FREQ.2_N | During the last 30 days, on how many days did you drink any alcoholic beverages, including beer, wine, or liquor? Please enter a numerical value. | Continuous |
| Alcohol quantity | ALC_QUANT_H | In the last three months, on the days you drink, about how many drinks do you have? Please enter a numerical value. | Continuous |
| Alcohol quantity | ALC_QUANT_N | On the days that you drink, about how many drinks do you have on the average day? By a drink, we mean the equivalent of a can of beer, a glass of wine, or a shot glass of hard liquor. Please enter a numerical value. | Continuous |
| Drinks per week | AVG_DRINKS_DAY_H | Product of alcohol consumption frequency (days drank per week) and drinks per day on days when alcohol was consumed | Continuous |
| Drinks per week | AVG_DRINKS_DAY_N | Product of alcohol consumption frequency (days drank per week) and drinks per day on days when alcohol was consumed | Continuous |

## Appendix 2: Harmonization Decisions

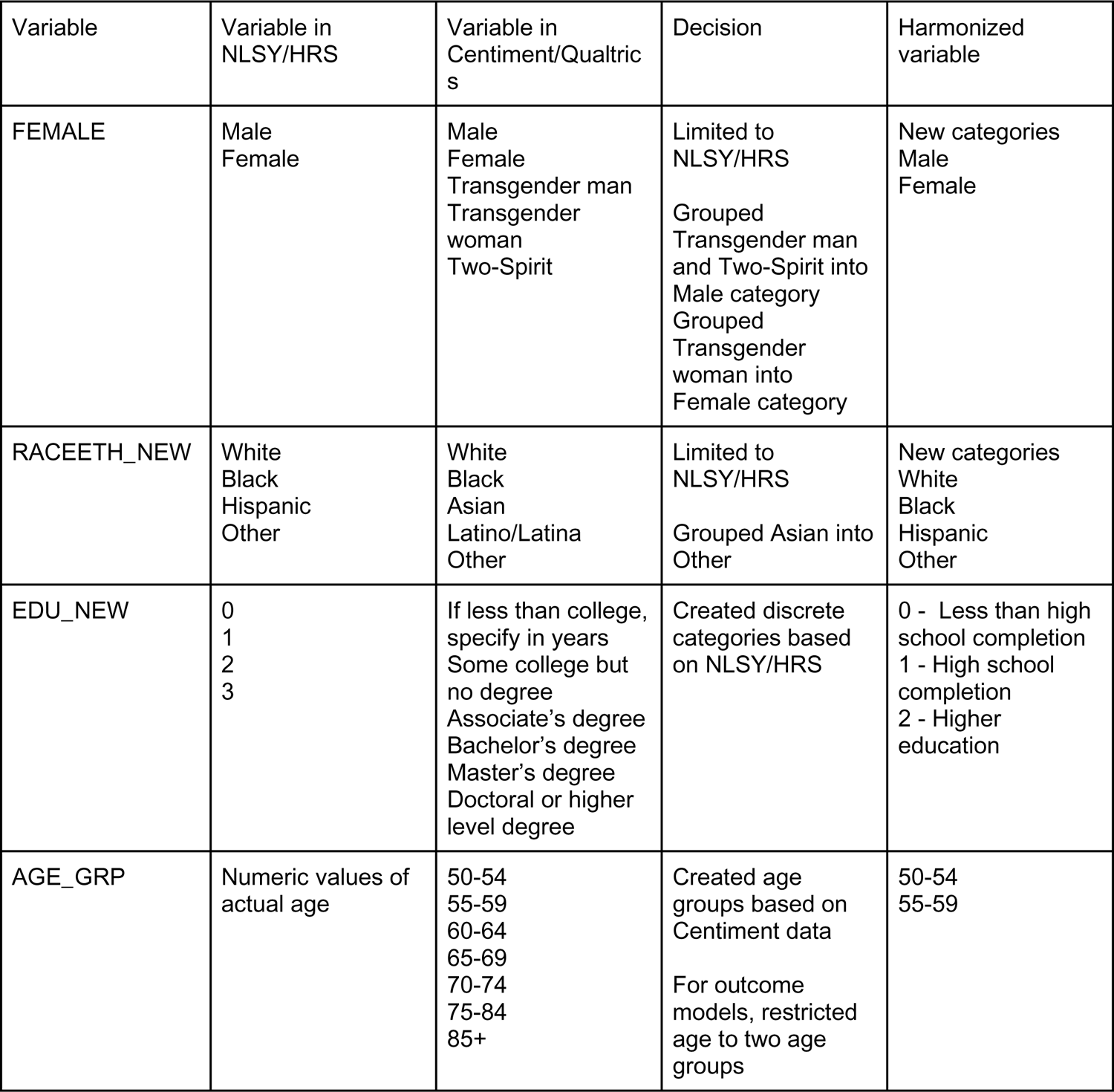

| Variable | Variable in NLSY/HRS | Variable in Centiment/Qualtrics | Decision | Harmonized variable |
| --- | --- | --- | --- | --- |
| FEMALE | Male<br>Female | Male<br>Female<br>Transgender man<br>Transgender woman<br>Two-Spirit | Limited to NLSY/HRS<br>Grouped Transgender man and Two-Spirit into Male category<br>Grouped Transgender woman into Female category | New categories<br>Male<br>Female |
| RACEETH_NEW | White<br>Black<br>Hispanic<br>Other | White<br>Black<br>Asian<br>Latino/Latina<br>Other | Limited to NLSY/HRS<br>Grouped Asian into Other | New categories<br>White<br>Black<br>Hispanic<br>Other |
| EDU_NEW | 0<br>1<br>2<br>3 | If less than college, specify in years<br>Some college but no degree<br>Associate's degree<br>Bachelor's degree<br>Master's degree<br>Doctoral or higher level degree | Created discrete categories based on NLSY/HRS | 0 - Less than high school completion<br>1 - High school completion<br>2 - Higher education |
| AGE_GRP | Numeric values of actual age | 50-54<br>55-59<br>60-64<br>65-69<br>70-74<br>75-84<br>85+ | Created age groups based on Centiment data<br>For outcome models, restricted age to two age groups | 50-54<br>55-59 |

## Appendix 3: Crosswalk scatterplots

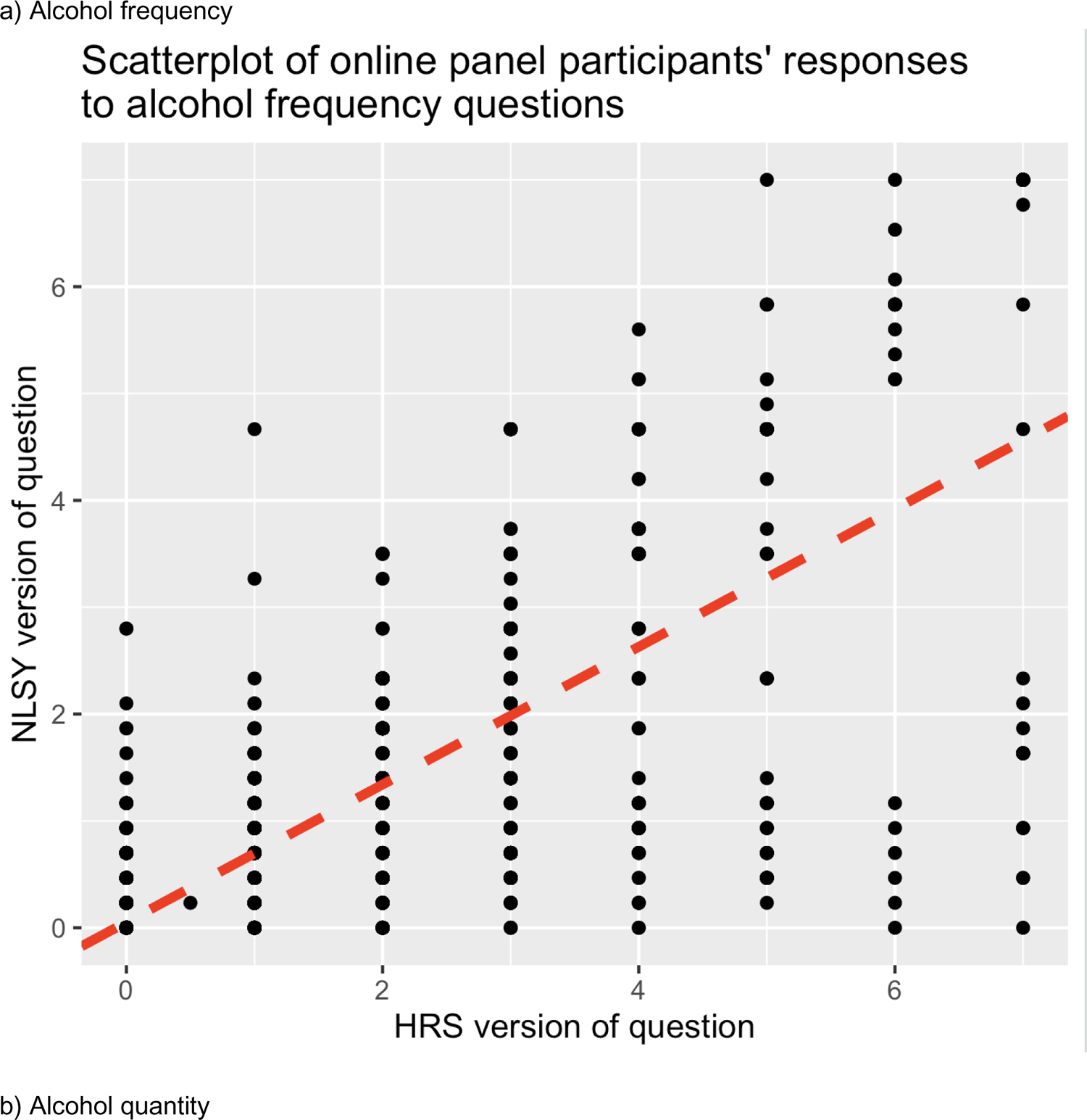
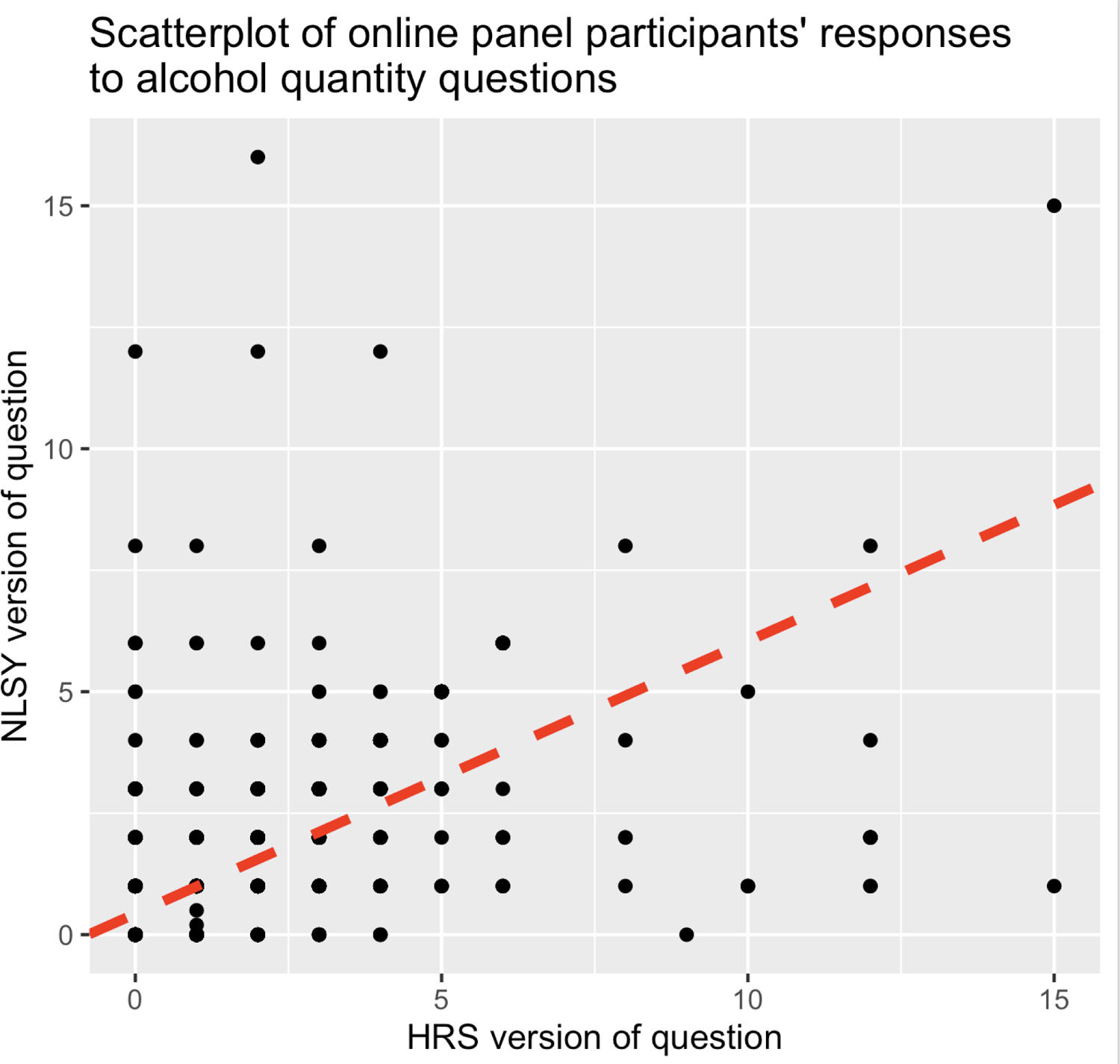
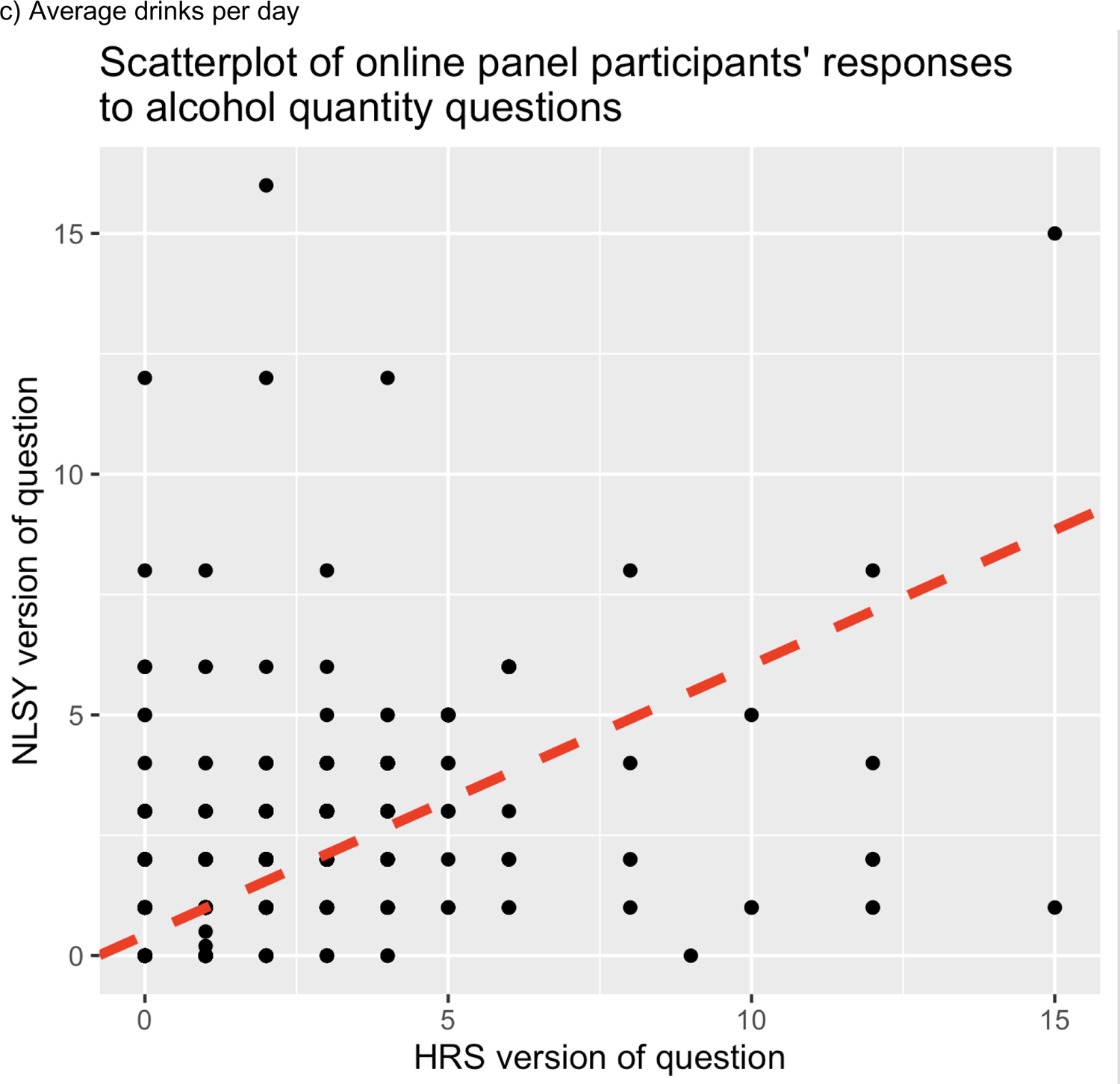

## Appendix 4: Full regression coefficients for general health models

| Domain | Data set | Question version | (Intercept) | Alcohol measure | Female | Race: Black | Race: Hispanic | Race: Other | Age group 55-59 | Education (completed high school) | Education (higher education) |
| --- | --- | --- | --- | --- | --- | --- | --- | --- | --- | --- | --- |
| Frequency | HRS | NLSY (imputed) | 2.121<br>[2.064,2.179] p<0.01 | -0.042 [-0.055,-0.029] p<0.01 | 0.089<br>[0.052,0.126] p<0.01 | 0.352<br>[0.307,0.397] p<0.01 | 0.312<br>[0.259,0.366] p<0.01 | 0.27<br>[0.193,0.346] p<0.01 | 0.096<br>[0.058,0.133] p<0.01 | -0.468 [-0.519,-0.417] p<0.01 | -0.934 [-0.993,-0.876] p<0.01 |
|  | NLSY | NLSY (original) | 1.965<br>[1.887,2.044] p<0.01 | -0.062 [-0.075,-0.049] p<0.01 | 0.114<br>[0.066,0.162] p<0.01 | 0.173<br>[0.116,0.231] p<0.01 | 0.081<br>[0.012,0.151] p=0.02 | 0.046 [-0.035,0.128] p=0.26 | 0.028 [-0.021,0.078] p=0.27 | -0.411 [-0.478,-0.343] p<0.01 | -0.82 [-0.904,-0.735] p<0.01 |
|  | HRS | HRS (original) | 2.141<br>[2.084,2.198] p<0.01 | -0.045 [-0.055,-0.036] p<0.01 | 0.082<br>[0.046,0.118] p<0.01 | 0.353<br>[0.309,0.396] p<0.01 | 0.309<br>[0.256,0.361] p<0.01 | 0.265<br>[0.189,0.341] p<0.01 | 0.096<br>[0.058,0.133] p<0.01 | -0.467 [-0.517,-0.416] p<0.01 | -0.932 [-0.989,-0.874] p<0.01 |
|  | NLSY | HRS (imputed) | 1.966<br>[1.883,2.048] p<0.01 | -0.051 [-0.065,-0.037] p<0.01 | 0.124<br>[0.075,0.173] p<0.01 | 0.192<br>[0.13,0.253] p<0.01 | 0.108<br>[0.036,0.18] p<0.01 | 0.048 [-0.034,0.13] p=0.25 | 0.016 [-0.036,0.068] p=0.55 | -0.408 [-0.477,-0.338] p<0.01 | -0.819 [-0.906,-0.732] p<0.01 |
| Quantity | HRS | NLSY (imputed) | 2.12<br>[2.056,2.185] p<0.01 | -0.03 [-0.049,-0.012] p<0.01 | 0.09<br>[0.052,0.128] p<0.01 | 0.367<br>[0.323,0.412] p<0.01 | 0.338<br>[0.285,0.392] p<0.01 | 0.271<br>[0.195,0.348] p<0.01 | 0.094<br>[0.057,0.132] p<0.01 | -0.477 [-0.529,-0.425] p<0.01 | -0.96 [-1.019,-0.901] p<0.01 |
|  | NLSY | NLSY (original) | 1.953<br>[1.873,2.034] p<0.01 | -0.041 [-0.055,-0.026] p<0.01 | 0.121<br>[0.072,0.169] p<0.01 | 0.19<br>[0.132,0.248] p<0.01 | 0.109<br>[0.04,0.179] p<0.01 | 0.055 [-0.027,0.137] p=0.19 | 0.019 [-0.031,0.069] p=0.45 | -0.424 [-0.492,-0.356] p<0.01 | -0.862 [-0.946,-0.778] p<0.01 |
|  | HRS | HRS (original) | 2.103<br>[2.045,2.161] p<0.01 | -0.02 [-0.029,-0.01] p<0.01 | 0.095<br>[0.058,0.131] p<0.01 | 0.369<br>[0.325,0.413] p<0.01 | 0.332<br>[0.279,0.385] p<0.01 | 0.277<br>[0.201,0.353] p<0.01 | 0.094<br>[0.057,0.132] p<0.01 | -0.471 [-0.522,-0.421] p<0.01 | -0.952 [-1.01,-0.894] p<0.01 |
|  | NLSY | HRS (imputed) | 1.943<br>[1.863,2.023] p<0.01 | -0.043 [-0.06,-0.026] p<0.01 | 0.127<br>[0.078,0.175] p<0.01 | 0.194<br>[0.135,0.252] p<0.01 | 0.111<br>[0.04,0.181] p<0.01 | 0.058 [-0.025,0.14] p=0.17 | 0.018 [-0.032,0.068] p=0.49 | -0.418 [-0.486,-0.35] p<0.01 | -0.847 [-0.932,-0.762] p<0.01 |
| Average daily drinking | HRS | NLSY (imputed) | 2.116<br>[2.057,2.176] p<0.01 | -0.019 [-0.027,-0.01] p<0.01 | 0.088<br>[0.051,0.126] p<0.01 | 0.36<br>[0.315,0.405] p<0.01 | 0.321<br>[0.267,0.374] p<0.01 | 0.271<br>[0.194,0.348] p<0.01 | 0.095<br>[0.057,0.133] p<0.01 | -0.47 [-0.521,-0.419] p<0.01 | -0.943 [-1.002,-0.885] p<0.01 |
|  | NLSY | NLSY (original) | 1.908<br>[1.83,1.986] p<0.01 | -0.006 [-0.01,-0.002] p<0.01 | 0.133<br>[0.085,0.181] p<0.01 | 0.198<br>[0.14,0.256] p<0.01 | 0.106<br>[0.036,0.176] p<0.01 | 0.059 [-0.023,0.142] p=0.16 | 0.023 [-0.027,0.073] p=0.37 | -0.424 [-0.492,-0.356] p<0.01 | -0.862 [-0.946,-0.777] p<0.01 |
|  | HRS | HRS (original) | 2.078<br>[2.021,2.136] p<0.01 | -0.002 [-0.004,0.001] p=0.15 | 0.107<br>[0.071,0.143] p<0.01 | 0.37<br>[0.326,0.414] p<0.01 | 0.329<br>[0.276,0.382] p<0.01 | 0.284<br>[0.208,0.36] p<0.01 | 0.095<br>[0.057,0.133] p<0.01 | -0.473 [-0.524,-0.422] p<0.01 | -0.95 [-1.008,-0.893] p<0.01 |
|  | NLSY | HRS (imputed) | 1.962<br>[1.877,2.047] p<0.01 | -0.02 [-0.027,-0.012] p<0.01 | 0.121<br>[0.071,0.171] p<0.01 | 0.194<br>[0.133,0.254] p<0.01 | 0.108<br>[0.037,0.178] p<0.01 | 0.05 [-0.033,0.133] p=0.24 | 0.012 [-0.041,0.065] p=0.66 | -0.416 [-0.485,-0.346] p<0.01 | -0.839 [-0.926,-0.751] p<0.01 |

Reference categories: race: white, age group: 50-54, education: "did not complete high school"

## Appendix 5: Full regression results from measurement models

| Domain | Independent alcohol variable question version | Dependent alcohol variable question version | Intercept | Coefficient on alcohol independent variable |
| --- | --- | --- | --- | --- |
| Frequency | HRS | NLSY | 0.07 [0.02,0.13] p=0.01 | 0.65 [0.63,0.67] p<0.01 |
|  | NLSY | HRS | 0.52 [0.46,0.59] p<0.01 | 0.94 [0.9,0.97] p<0.01 |
| Quantity | HRS | NLSY | 0.51 [0.44,0.58] p<0.01 | 0.55 [0.52,0.57] p<0.01 |
|  | NLSY | HRS | 0.42 [0.33,0.51] p<0.01 | 0.84 [0.8,0.87] p<0.01 |
| Average daily drinking | HRS | NLSY | 0.25 [0.15,0.34] p<0.01 | 0.53 [0.51,0.55] p<0.01 |
|  | NLSY | HRS | 0.85 [0.73,0.98] p<0.01 | 0.95 [0.91,0.99] p<0.01 |

## Notes

### Competing Interest Statement

The authors have declared no competing interest.

### Funding Statement

This study was funded by NIH-NIA grants #R01AG072681.

### Author Declarations

IRB of the University of California, San Fransisco, waived ethical approval for this work.

